# Efficacy and safety of Tenofovir-based regimen in elderly people living with HIV in sub-Saharan Africa: A systematic review and meta-analysis

**DOI:** 10.1101/2022.11.01.22281791

**Authors:** Folahanmi T. Akinsolu, Sabdat Ekama, Adesola Z. Musa, David Oladele, Esther Ohihion, Priscilla Ezemelue, Osuolale K. Adewale, Flora Davies-Bolorunduro, Emelda Chukwu, Muinah Fowora, Sola Ajibaye, Sam Amoo, Ifeoma Idigbe, Toyosi Raheem, Ebiere Herbertson, Aisha Gambari, Abideen Salako, Mobolaji Olagunju, Titi Gbaja-biamila, Tosin Odubela, Pascal Ezeobi, Jane Okwuzu, Agatha N. David, Nkiruka Odunukwe, Ifeanyichukwu Ezebialu, George U. Eleje, Oliver C. Ezechi

**Affiliations:** Nigerian Institute of Medical Research, Lagos; Lead City University, Ibadan; Chukemuemeka Odumegwu Ojukwu University, Awka Campus, Nigeria; Nnamdi Azikiwe University, Awka, Nigeria

**Author notes:** Corresponding Author: Nigerian Institute of Medical Research, Lagos, and Lead City University, Ibadan. These authors contributed equally to this work (Folahanmi T. Akinsolu, Sabdat Ekama, Adesola Z. Musa, David Oladele, Esther Ohihion, Priscilla Ezemelue, Nkiruka Odunukwe, Ifeanyichukwu Ezebialu, George U. Eleje, Oliver C. Ezechi). These authors also contributed equally to this work (Osuolale K. Adewale, Flora Davies-Bolorunduro, Emelda Chukwu, Muinah Fowora, Sola Ajibaye, Sam Amoo, Ifeoma Idigbe, Toyosi Raheem, Ebiere Herbertson, Aisha Gambari, Abideen Salako, Mobolaji Olagunju, Titi Gbaja-biamila, Tosin Odubela, Pascal Ezeobi, Jane Okwuzu, Agatha N. David).

**Keywords:** Geriatrics, kidney disease, osteoporosis, randomized trial, Viread

## Abstract

**Objective:** The study examined whether the benefit and adverse effects of the tenofovir-based highly active antiretroviral therapy (HAART) regimen outweigh the non-tenofovir-based regimen in the elderly population.

**Methods:** We searched PubMed, EMBASE, Cochrane Central Library, Google Scholar, and some hand searches on September 21, 2021, to identify eligible studies. Only randomized control trials on elderly HIV-positive patients on Tenofovir-based regimens living in sub-Saharan Africa were included. Studies on pregnant women or prophylactic tenofovir were excluded. The primary outcomes are viral suppression, mortality, and anemia. Two reviewers independently selected the studies, extracted data, and assessed the risk of bias. We analyzed using risk ratio, with 95% confidence intervals. A fixed effect model along with an assessment of heterogeneity was done for meta-analysis.

**Results:** Four studies with a total of 263 participants were included. Our meta-analysis shows that there was no difference between participants on a tenofovir-based regimen and the non-tenofovir-based regimen in terms of viral suppression, mortality, anemia, and hypertension.

Our meta-analysis shows that there was no difference between participants on tenofovir based regimen and non-tenofovir based regimen in terms of viral suppression (RR=1.96, 95% CI (1.42 -2.70; I^2^=0%, 4 trials, 263 participants, very low certainty of evidence), mortality (RR = 2.90, 95% CI (0.12 – 69.87, I^2^=Not estimated, a trial, 120 participants, very low certainty of evidence), anaemia (RR=1.61, 95% CI (1.02-2.90; I^2^ = 0.16, 2 trials, 154 participants, very low certainty of evidence), hypertension RR = 0.76, 95% CI (0.44-1.31) I^2^ = Not estimated, a trial, 34 participants, low-certainty of evidence). None of the trials reported the incidence of chronic kidney disease and bone demineralization.

**Conclusion:** There was very-low certainty evidence that no difference exists between the tenofovir-based HAART regimen and the non-tenofovir-based regimen in terms of benefits and short-term adverse outcomes. Well-designed randomized clinical trials are needed with a focus on long-term adverse effects.

**ARTICLE SUMMARY:** *Strengths and limitations of the study:*

- Tenofovir-based high-active antiretroviral therapy (HAART) is one of the preferred first-line therapies in the management of HIV-1 infection.
- The study demonstrates evidence and represents the first comprehensive and up-to-date systematic review and meta-analysis on the effects of the tenofovir-based regimen on elderly patients concerning chronic kidney disease, changes in bone mineral density, number of deaths, viral suppression, hypertension, anemia, and adherence levels of ≥ 95%.
- The clinical implications of the study indicate there is an urgent need for evaluation of the effect of long-term use of tenofovir-based regimens among elderly people living with HIV/AIDS due to the paucity of data on the use of tenofovir-based HAART regimens.
- The study limitations are the cumulative sample size of the four studies was small with only one study reporting on mortality outcome; there was also a lack of statistical power for the clinical outcomes observed; there was no information on gender disaggregation to consider conducting a sub-group analysis; and there was a great amount of heterogeneity in one of the studies, which was accounted for by using a random effects model but has the same value in a fixed effect model.

## INTRODUCTION

The Human Immunodeficiency Virus/Acquired immunodeficiency syndrome (HIV/AIDS) infection pandemic continues to have devastating health effects globally, with approximately 75.7 million people of which over 95% are adults, and cumulative mortality of 32.7 million ^1, 2^. Sub-Saharan Africa (SSA) accounts for two-thirds of the global burden^1, 2^. With the high uptake/success of high active antiretroviral therapy (HAART), the age of people living with HIV/AIDS (PLWHA) has been rising steadily resulting in a huge population of elderly people living with HIV. WHO defines these elderly people as aged 60 years and more^3^, which age criteria has recently been moved to 65 years and above in Europe and America^2^.

Tenofovir is a nucleotide reverse transcriptase inhibitor used in combination with other classes of antiretroviral drugs in a standard HAART combination. Tenofovir-based HAART is one of the preferred first-line therapies in the management of HIV-1 infection^4^. Tenofovir disoproxil fumarate (TDF) and Tenofovir Alafenamide (TAF) are prodrugs of Tenofovir. TDF is activated by initial hydrolysis to tenofovir and then phosphorylation to tenofovir diphosphate. It is excreted through glomerular filtration and active tubular secretion. Tenofovir diphosphate inhibits the activity of HIV-1 reverse transcriptase by competing with the natural substrate, deoxyadenosine 5’-triphosphate, with a resultantly DNA chain termination. TAF is a novel prodrug with improved efficacy and reduced renal toxicity potentials compared to TDF^5^. Tenofovir disoproxil fumarate is administered as a once-daily 300 mg tablet commonly co-formulated with emtricitabine. Common side effects associated with tenofovir are renal impairment and bone mineral density loss^4^.

The advent of antiretroviral therapy for the treatment of HIV/AIDS infection is one of the greatest breakthroughs of modern medicine. HIV-infected patients in this wake shifted from a lifespan in years to decades^6^. The initial regimens utilized were complex and associated with significant short-term and long-term toxicity, however current regimens are generally easier to administer, safer, and better tolerated of which Tenofovir is a classic example. In this generally acceptable mode where the reality is that of control of the HIV epidemic, added to the fact of being able to achieve, and indefinitely maintain control of HIV replication in the vast majority of patients, there is a need to determine the effect of these treatment regimens.

Despite the unquestioned success, combination antiretroviral therapy does not fully restore health. For reasons that remain poorly defined, long-term treated HIV-infected persons have an expected life span that is substantially shorter than that of their HIV-uninfected peers^7–10^. This shortened life span is largely due to an increased risk of some “non-AIDS” complications, encompassing heart disease, cancer, liver disease, kidney disease, bone disease, and neurocognitive decline^8^ many of which complications are similar to that observed generally among the elderly. In addition, the fact of these diseases are degenerative in nature will affect the quality of life and functionality.

More specifically, the use of a tenofovir-based regimen over a long period among HIV-positive individuals should be assessed for its long-term effect especially among the elderly because of prolonged antiretroviral drug use considering potential renal toxicities associated with the drug and the tendency for polypharmacy among these cohorts of individuals. As well as the excess burden of comorbid diseases among this cohort of individuals^11^. The fact of antiretroviral treatment toxicity is well established is not in doubt, and there is epidemiologic, clinical, and pathogenesis data supporting the concept that antiretroviral-treated HIV-infected persons are at higher-than-normal risk for certain age-associated diseases^11^. Therefore, we systematically reviewed the published literature to examine the number of elderly patients with viral suppression, chronic kidney disease, changes in bone mineral density, number of deaths, anemia, hypertension, and adherence levels of ≥ 95% while on a tenofovir-based HAART regimen.

## METHODS

### Protocol Registration

We did a systematic review of studies on the efficacy and safety of the Tenofovir-based regimens in elderly people living with HIV in sub-Saharan Africa. The review was registered prospectively with PROSPERO (CRD42021278695) and reported according to PRISMA 2020 guidelines^12^.

### Search Strategy

An extensive online search was conducted using PubMed, Google Scholar, Central Cochrane Library, and some hand searches from inception to date. Medical Subject Headings (MeSH) and free text words were combined using Boolean operators “OR” and “AND.” The reference lists of included studies were screened to identify additional publications.

In this systematic review, we searched for studies that reported on the efficacy and safety of the Tenofovir-based regimens in elderly people living with HIV in sub-Saharan Africa. The search was done from inception to date. The searches were done with no language restrictions on 25 September 2021, in PubMed, Google Scholar, and Central Cochrane Library. The search terms included were middle-aged [Mh] OR Aged [Mh] AND AIDS [Mh] OR HIV-1 [Mh] OR HIV-2 [Mh] OR Acquired Immune Deficiency Syndrome Virus” [Mh] AND Tenofovir[mh] AND RCT OR randomized clinical trial OR randomized clinical trial AND sub-Saharan Africa (See S1). Searches were tailored to each database. Reference lists were screened for additional sources. The search focused on published medical literature as well as gray literature.

All searches were based on relevant titles, abstracts, or keywords. We excluded abstracts for poster presentations where no full details were not available. We also examined the reference lists of the retrieved articles and reviewed papers to identify any studies missed by the initial search.

### Eligibility criteria

We included randomized controlled trials in which a tenofovir-based regimen was administered as an intervention to the study participants to monitor treatment outcomes in comparison to the non-tenofovir-based regimen. The study population was elderly HIV-positive patients aged 50 years and above on antiretroviral therapy specifically a tenofovir-based regimen in comparison with a non-tenofovir-based regimen.

### Study Population

The study population was defined as elderly/aged people living with HIV in sub-Saharan Africa.

### Study Area

Only studies conducted in sub-Saharan Africa were eligible for inclusion.

### Language

Studies reported were not restricted to the English language.

### Publication condition

Studies that meet the eligibility criteria were included regardless of their publication status (published, unpublished, or grey literature).

### Information Sources

Four major electronic databases were screened from inception until September 21, 2021. These databases include PubMed, Google Scholar, Central Cochrane Library, and EMBASE. Additionally, the reference list of the included RCTs and recent reviews were manually screened for additional important RCTs that could have been overlooked. There were no restrictions on language or year of publication.

### Main Outcomes

The main outcome measure was the number of participants with viral suppression, the number of mortalities, and the number of participants with anemia. The secondary outcomes measured include the proportions of participants with (i) hypertension (ii) chronic kidney disease; (iii) changes in bone mineral density/osteoporosis/bone loss, and (iv) drug adherence levels of ≥ 95%.

### Study Selection

All titles and abstracts identified as potentially relevant were assessed. Two reviewers (SE and DO) independently searched and selected the studies to be included. A third reviewer (IE) resolved issues with discrepancies in study selection. Duplicated studies were removed from search results.

### Data Extraction

Study Characteristics including the settings, interventions, comparisons, and outcome measures were extracted by ZA and GE. Data for the meta-analysis were extracted as raw data. The data for all included studies was collected using a formatted spreadsheet according to the “PICO” format (patient, intervention, comparison, and outcome) of the published results.

### Analysis and Quality Assessment

We used a fixed effect for the meta-analysis (RevMan Version 5.4.1)^13^. Risk ratios were calculated for available outcomes and reported with 95% confidence intervals (CI). Statistical heterogeneity was evaluated using the *I*^*2*^ index.

The risk of bias for each study was determined based on statements relating to random sequence generation, allocation concealment, blinding of participants and personnel, outcome assessment, incomplete outcome data, and selective reporting. The GRADE approach provided the framework for rating the certainty of the evidence for each outcome. Concerns with risk of bias, inconsistency, indirectness, imprecision, and publication bias lowered certainty. We considered the bodies of evidence from RCTs. Each property was ranked as “very low risk”, or “low risk”, according to the quality of the GRADE profile tool and presented as a summary of findings.

### Subgroup analysis and investigation of heterogeneity

#### Sensitivity analysis

We carried out a sensitivity analysis to explore the effects of trial quality assessed by allocation concealment and another risk of bias components, by omitting studies rated as “high risk of bias” for these components. We restricted this to the primary outcomes.

#### Patient and Public Involvement

No patient was involved in this study.

## RESULTS

### Description of studies

Results of the search. The search strategy yielded a total of 757 potentially relevant studies (see Figure 1). When the search was exported into the endnote and duplicates were removed, there were 96 unique records. Two authors (FT and GE) independently read the titles and abstracts and excluded 92 studies because they did not meet the inclusion criteria. Four studies met the inclusion criteria as shown in Figure 1, Table 1^14-17^. The four studies were carried out in health facilities within low- and middle-income countries. Data were extracted from a total number of 263 persons living with HIV/AIDS receiving a tenofovir-based regimen in Lesotho, Senegal, Burkina Faso, and Cameroon. Table 2 presents the main characteristics of the 4 included studies ^14-17^.

**Figure 1.**
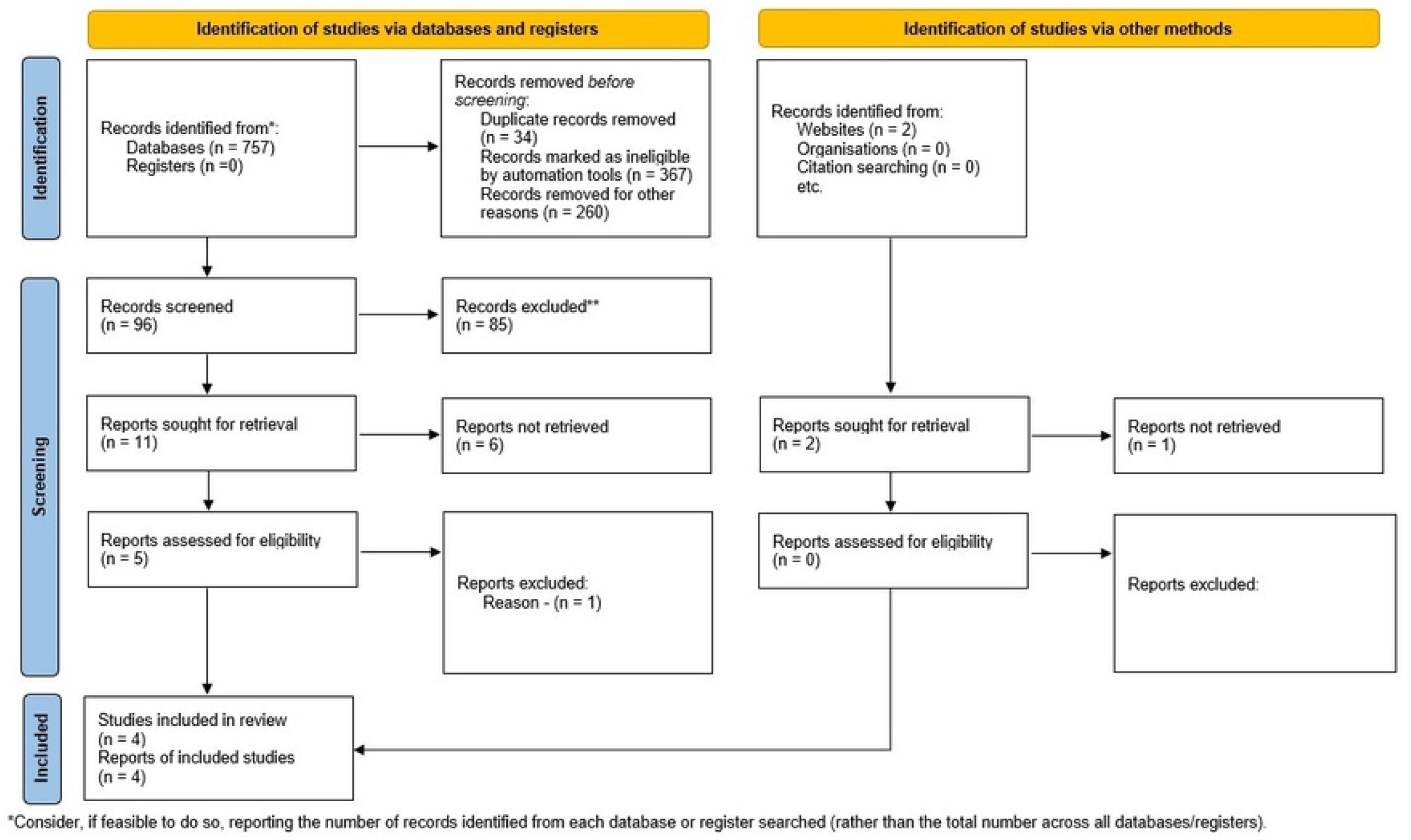
Flow chart of the literature search. The diagram shows the different phases of the systematic review.

**Table 1:** Studies included in the meta-analysis.

| Study (year) | N | Study population | Country Study | Assessment | Intervention | Comparison | Primary outcome measure | Secondary outcome measure |
| --- | --- | --- | --- | --- | --- | --- | --- | --- |
| Amstutz 2020<sup>14</sup> | 80 | An individual living with HIV | Lesotho | Assessment of patients with sustained low-level HIV-positive on first-line ART benefit from a switch to second-line treatment. | Non-nucleoside reverse transcriptase inhibitor (NNRTI)-based first-line ART Tenofovir/ emtricitabine + lopinavir/ritonavir or abacavir + didanosine + lopinavir/ritonavir or tenofovir/emtricitabine + darunavir/ritonavir (TDF/FTC + LPVr, ABC + ddI + LPVr or TDF/FTC + DRVr) | first-line ART containing non-nucleoside reverse transcriptase inhibitors (NNRTI) | VL<50 copies/mL at 36 weeks after randomization; | VL<100, <200, <400, <1000 copies/mL at 36 weeks; VL<50 copies/mL at 24 weeks; and VL≥ 50 copies/mL at 24 weeks and 36 weeks<br>Changes in body weight<br>Hemoglobin<br>CD4 count<br>Blood lipids at 36 weeks |
| Ba 2018<sup>15</sup> | 30 | HIV-infected; ART-naive adults with WHO stage 3 or 4 disease or CD4 counts <750 cells/μL | Senegal | Effectiveness of drugs in HIV-infected individuals | Single-tablet regimen containing elvitegravir, cobicistat, emtricitabine, and tenofovir disoproxil fumarate (E/C/F/TDF) | cotrimoxazole | clinical and immunovirological outcomes as well as adverse events | Changes in HIV pVL, CD4 cell counts, adverse events, all-cause mortality, and loss of follow-up |
| Kabore 2017<sup>16</sup> | 34 | HIV- positive patients | Burkina Faso, Cameroon, and Senegal | Assess the change in bone quality using quantitative ultrasound (QUS) over 96 weeks of follow-up | TDF/FTC/LPVr, ABC/ddI/LPVr or TDF/FTC/DRVr. | ABC+ddI +LPVr | The morphological changes and metabolic disorders (i.e., metabolic syndrome and cardiovascular and bone changes) in HIV-positive patients who failed first-line antiretroviral treatment, and their evolution on second-line ART. |  |
| Landman 2014<sup>17</sup> | 119 | HIV positive Individuals in Senegal, or Cameroon, ARV, and CD4 T-cell count >50 cells/mm<sup>3</sup> | Cameroon and Senegal | Determine appropriate tenofovir-based regimens meriting evaluation in large-scale randomized trials | tenofovir/emtricitabine/nevirapine (group 1), tenofovir/lopinavir/ritonavir (group 2), tenofovir/emtricitabine/zidovudine (group 3) and tenofovir/emtricitabine/efavirenz (group 4) |  | Virological efficacy (HIV-1 RNA<50 copies/ml at week 16 in ≥50% of patients) | The proportion of patients with either HIV viral load ≥800 copies/ml at week 16 or ≥100 copies/ml from week 24 to week 96, increase of CD4+ T-cell count from baseline, and proportion of clinical and laboratory adverse events. |

**Table 2:**
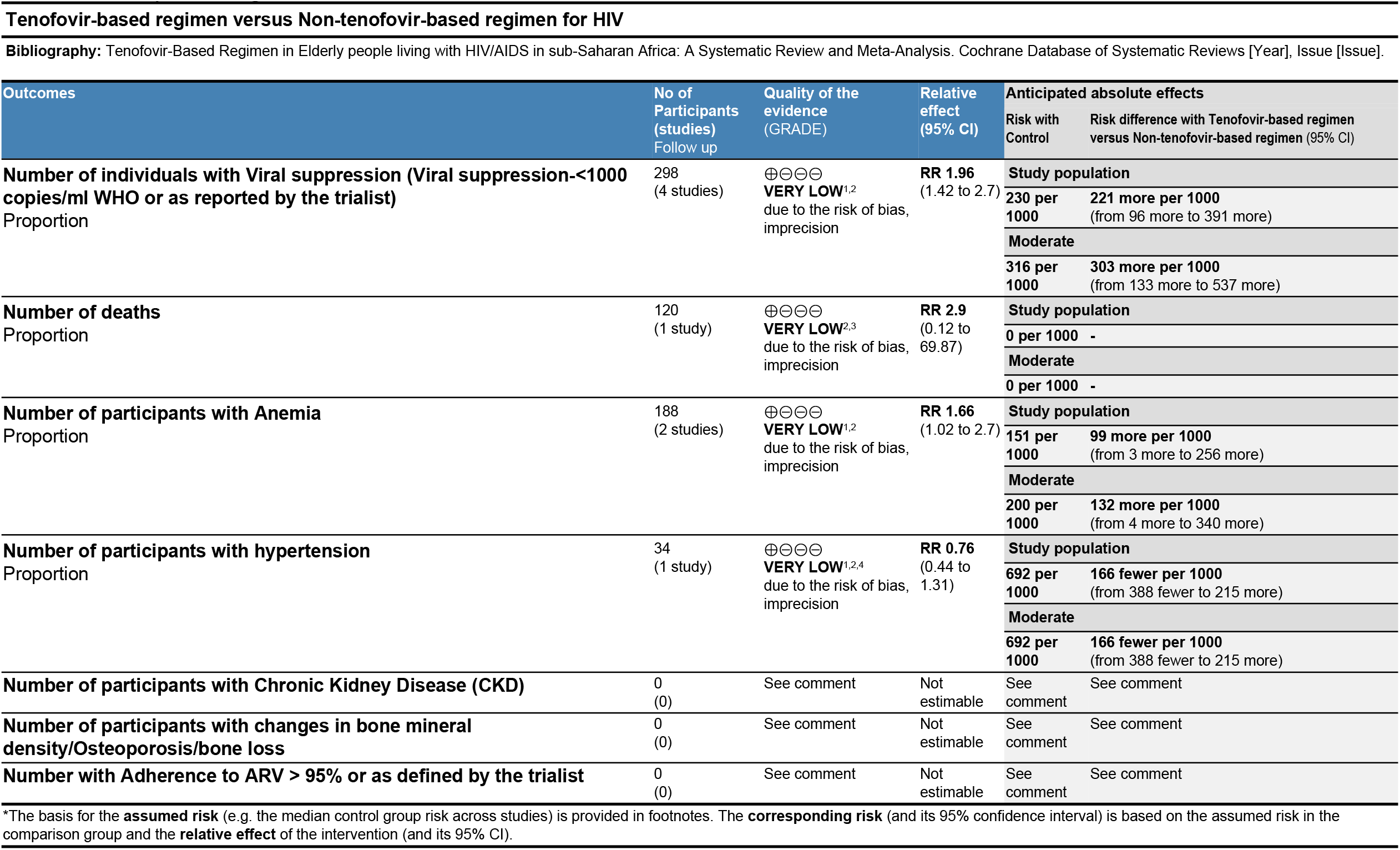
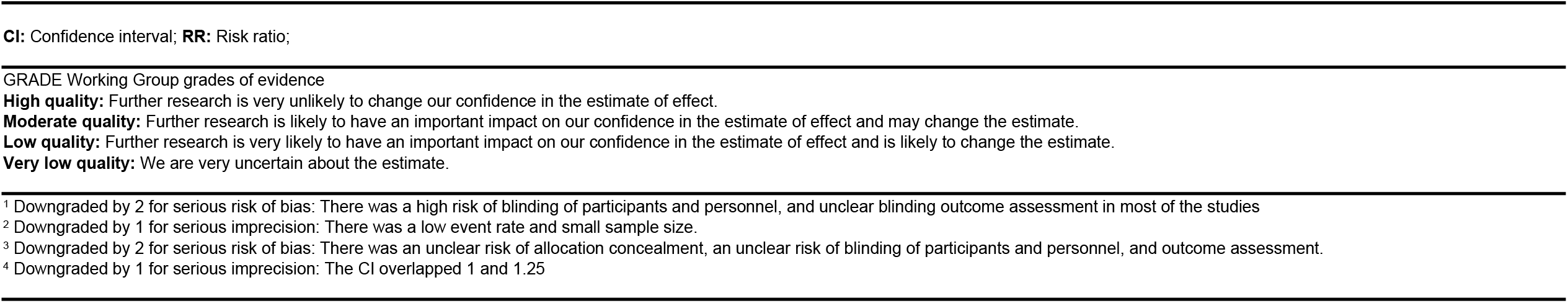
Summary of Findings.

### Data collection and analysis

#### Viral suppression

The four included studies reported on viral suppression outcomes which shows that there is no significant difference between the tenofovir-based regimen and the non-tenofovir-based regimen (RR = 1.96, 95% CI (1.42 – 2.70); 4 trials, very low certainty of the evidence)^14-17^. (See Figure 2a)

**Figure 2.**
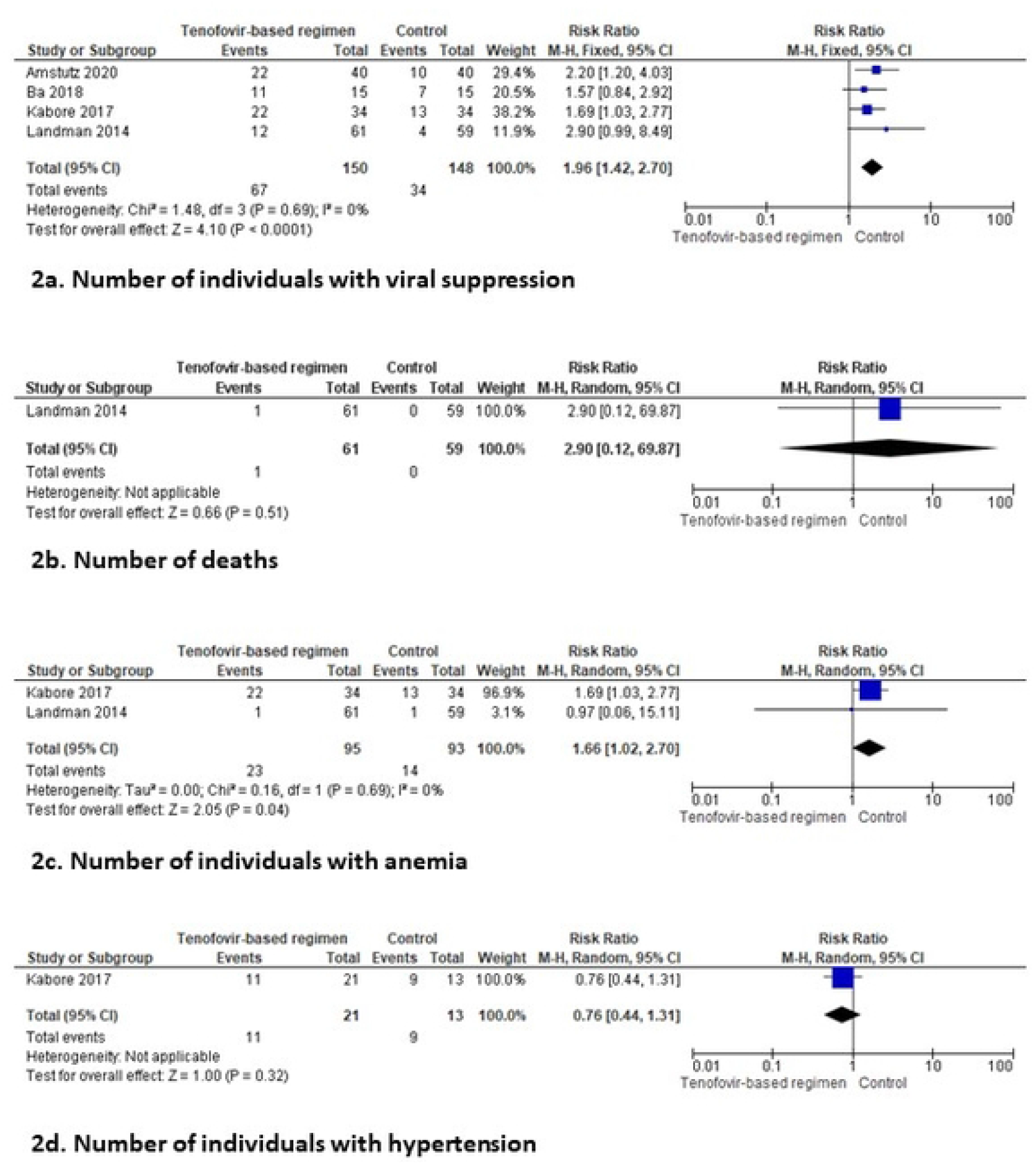
Forest plot of the observed effect of tenofovir-based regimen in the included studies. Effect of tenofovir-based regimen on (A) number of individuals with viral suppression, (B) number of deaths, (C) number of individuals with anemia, and (D) number of individuals with hypertension.

#### Mortality

A single study reported on the mortality among elderly people living with HIV in sub-Saharan Africa on the TDF regimen^17^. The meta-analysis shows no observed heterogeneity for mortality (RR = 2.90, 95% CI (0.12 – 69.87), *I*^*2*^ = Not estimated, 1 trial, very low certainty of evidence). (See Figure 2b)

#### Anemia

Two of the included studies reported on some individuals with anemia. The studies showed no evidence of statistical heterogeneity (*I*^*2*^ = 0%; p > 0.05)^16, 17^. The included studies likely have a relatively small sample size (RR=1.66, 95% CI (1.02 – 2.70); *I*^*2*^ = Not estimated, 2 trials, very low certainty of the evidence). (See Figure 2c)

#### Hypertension

A single study reported hypertension among elderly people living with HIV in sub-Saharan Africa on the TDF regimen^16^. The meta-analysis shows no observed heterogeneity for hypertension (RR = 0.76, 95% CI (0.44 – 1.31), *I*^*2*^ = Not estimated, 1 trial, very low certainty of evidence). (See Figure 2d)

Each study was also assessed for the risk of bias (shown in Figure 3 and Table 2). None of the included studies reported on the blinding of outcome assessment. Two of the studies discussed in detail the process of their random sequence generation about selection bias. It is also important to note that all the included studies have a low risk for attrition bias (See Appendix II).

**Figure 3.**
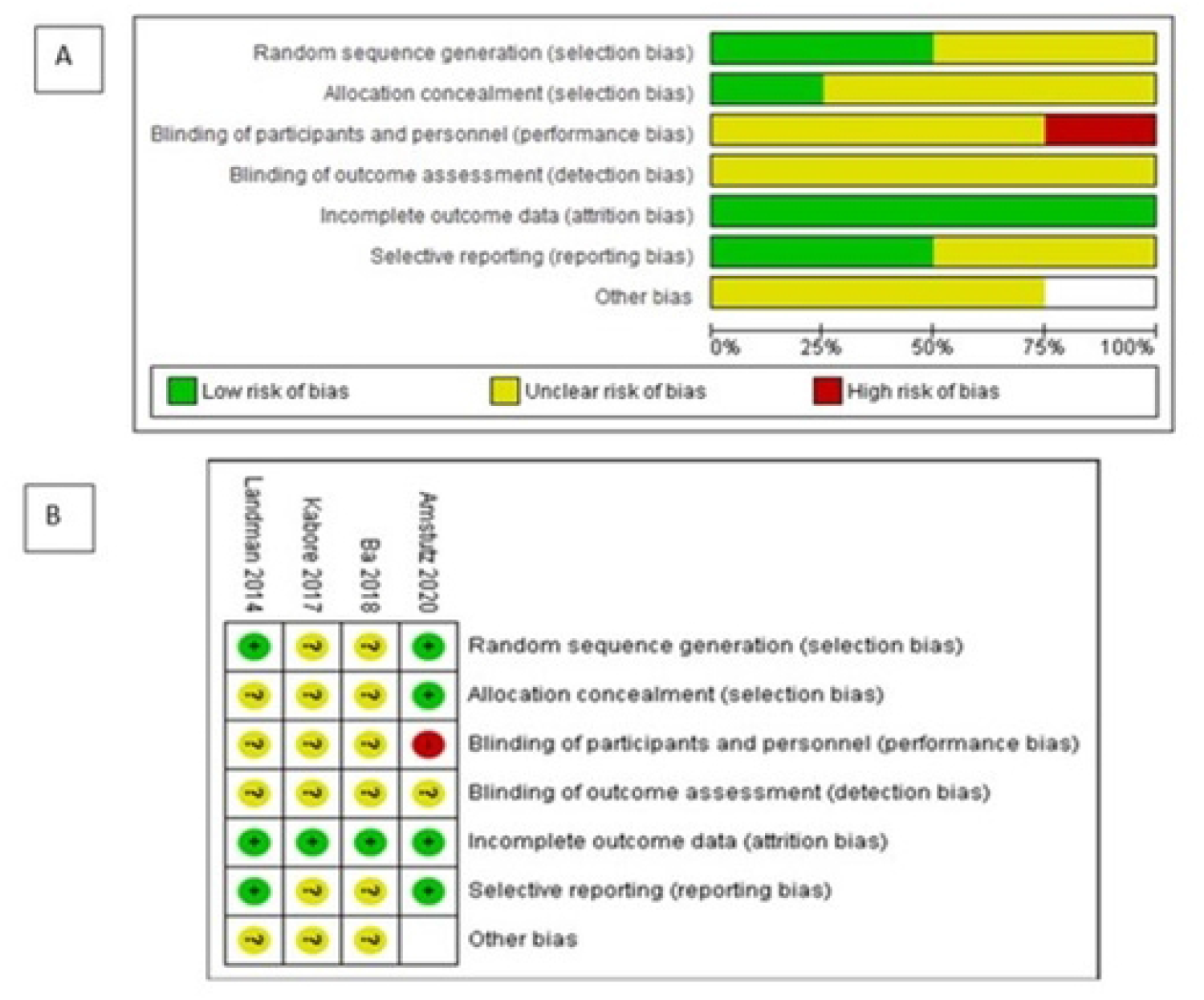
Risk of bias assessment. (A) Risk of bias graph and (B) Risk of bias summary.

## Discussion

In this systematic review and meta-analysis, four studies were assessed that reported the treatment outcome of the tenofovir-based highly active antiretroviral therapy (HAART) regimen among elderly persons living with HIV/AIDS in sub-Saharan Africa. The studies included identified the use of the nucleotide reverse transcriptase inhibitor (tenofovir) as the backbone for the HAART regimen, and it shows no difference in terms of the outcomes observed compared to the non-tenofovir-based regimen. None of the studies reported the incidence of chronic kidney disease, bone demineralization, and adherence levels.

To the best of our knowledge, this is the first systematic review and meta-analysis on the use of a tenofovir-based regimen among the elderly population living with HIV/AIDS in sub-Saharan Africa. We detected a meta-analysis conducted by Chen et al., 2021 which focused only adult population^18^. In Chen et al., study, seven published RCTs were included with a total of 3547 participants^18^. The study shows no significant difference between viral suppression and efficacy outcome of the tenofovir-based regimen and non-tenofovir-based regimen [odds ratio (OR) 1.01 (95% CI 0.79, 1.30)]. Our study meta-analysis (RR=1.96, 95% CI (1.42 – 2.70; *I*^*2*^=0%), is in tandem with their findings on viral suppression Bictegravir/Emtricitabine/TAF had comparable safety profiles of control regimens.

In this review, none of the trials reported the incidence of chronic kidney disease, bone demineralization, and adherence level. A previous systematic review by Mtisi et al^19^, revealed the development of kidney disease in HIV-positive African patients on a tenofovir-disoproxil fumarate-containing antiretroviral regimen. The identified studies in Africans reported statistically significant renal function decline associated with tenofovir-disoproxil-fumarate use but the clinical significance of this effect was not enough to contraindicate its continued use in ART regimens. Mtisi et al^19^ further concluded that patients are at greater risk if they have pre-existing renal disease and are more advanced in age. However, studies included in the Mtisi et al review were not limited to RCTs but also observational studies. Also, in another study by Tan et al^20^, TDF-related renal impairment remained rare in HIV-positive Chinese patients with a median age of 29 years who had no comorbidities. Lower weight and duration of ART were associated with decreased renal function. Another study by Yilma et al^21^ reported that renal function remained stable with no difference between HIV patients treated with TDF or non-TDF non-nucleoside reverse transcriptase inhibitor-based ART regimen over 12 months. However, older HIV patients and those with unsuppressed viral loads deserve special focus on renal monitoring. This lack of studies eligible in the present review is of concern since ours is limited to randomized control trials and the elderly population.

This study shows that there is no significant difference between participants on the tenofovir-based regimen and the non-tenofovir-based regimen in terms of mortality (RR = 2.90, 95% CI (0.12 – 69.87), anemia (RR=1.61, 95% CI (1.02-2.70) and hypertension (RR = 0.76, 95% CI (0.44-1.31). This is akin to a previous study by Mulenga et al^22^ that reported that patients receiving TDF-containing HAART, despite pre-existing renal disease, did not experience worse renal outcomes or increased mortality compared with those taking other regimens. The same can be said of anemia and hypertension. Anemia can result from chronic renal disease while hypertension could be due to the elderly population of the study.

The clinical implications of the study indicate there is an urgent need for evaluation of the effect of long-term use of tenofovir-based regimens among elderly people living with HIV/AIDS due to the paucity of data on the use of tenofovir-based HAART regimens. The included studies have unclear blinding of outcome assessment while three of the studies did not discuss their participants’ allocation concealment. This calls for more robust and adequately conducted RCTs in the future.

This study has limitations, although, the data reported in this study meta-analysis show some beneficial effects of a tenofovir-based regimen on viral suppression, risk of anemia, deaths, and high blood pressure. Firstly, the cumulative sample size of the four studies was small with only one study reporting on mortality outcomes. Secondly, there was also a lack of statistical power for the clinical outcomes observed. Thirdly, there was no information on gender disaggregation to consider conducting a sub-group analysis. Fourthly, there was a great amount of heterogeneity in one of the studies, which was accounted for by using a random effects model but has the same value in a fixed effect model.

Despite these limitations, the present study represents the first comprehensive and up-to-date systematic review and meta-analysis on the effects of a tenofovir-based regimen on elderly patients concerning chronic kidney disease, changes in bone mineral density, number of deaths, viral suppression, hypertension, anemia and adherence levels of ≥ 95%. A thorough literature search, and assessment of a different tenofovir-based highly active antiretroviral therapy (HAART) regimen–based clinical outcomes represent another main strength of the present systematic review and meta-analysis.

## Conclusion

There is very low-certainty evidence on the benefits or harm of a tenofovir-based regimen over a non-tenofovir-based regimen on elderly HIV-positive individuals. The pooled data from the studies suggest that there was no difference between the tenofovir-based HAART regimen and the non-tenofovir-based regimen based on viral suppression, mortality, anemia, and hypertension. No data was available on the incidence of chronic kidney disease, bone demineralization, and adherence levels. Well-designed randomized clinical trials are needed on the use of tenofovir-based regimens among the HIV-positive elderly population with a focus on long-term adverse effects.

## Data Availability

All relevant data are within the manuscript and its Supporting Information files.

## Contributors

Conceptualization: Oliver Ezechi

Data curation: All authors

Formal analysis: All authors

Funding acquisition: All authors

Investigation: All authors

Methodology: All authors

Project administration: All authors

Resources: Oliver Ezechi

Software: George Eleje

Supervision: George Eleje, and Ifeanyichukwu Ezebialu

Validation: All authors

Visualization: All authors

Writing–original draft: All authors

Writing– review & editing: All authors

## Data Sharing

All datasets generated and analyzed, including the study protocol, search strategy, list of the included and excluded studies, data extracted, analysis plans, and quality assessment, are available in the Article and upon request from the corresponding author.

## Acknowledgments

The authors would like to thank Moriam Chibuzor of Cochrane Nigeria for her role in developing the search strategy and electronic searching of databases.

## Ethical approval

Ethical approval was not required for this systematic review as the research was based on information retrieved from published studies.

## Supplementary materials

**S1 – Search strategy**

